# The blood microbiome map of one million east asian whole-genome sequences from plasma cell-free DNA

**DOI:** 10.1101/2025.11.05.25339628

**Authors:** Zhixu Qiu, Kaixuan Duan, Shaohui Qian, Linfeng Yang, Shuo Li, Zhe Lin, Hankui Liu, Meng Wu, Sijie He, Yanan Niu, Cuixin Qiang, Pu Qin, Zhirong Li, Jing Yang, Weigang Wang, Yunfang Wang, Defeng Tao, Chengbin Yan, Shuyang Gao, Jianguo Zhang, Xiangdong Xu, Lijian Zhao, Jing Liu, Jianhong Zhao

## Abstract

Microbial cell-free DNA (mcfDNA) analysis offers a novel approach for monitoring blood-borne viral infections, with prior studies highlighting its clinical utility in viral lineage characterization. However, existing research predominantly employs cross-sectional designs, lacking systematic investigation into spatiotemporal viral dynamics and host genetic regulation. Leveraging non-invasive prenatal testing (NIPT) data from 1,023,651 pregnant women in Hebei Province (2019-2022), we constructed the largest spatiotemporal virome map during pregnancy. Analysis revealed higher viral diversity and load in plain regions compared to mountainous/coastal areas. Temporally, most viruses exhibited annual prevalence increases with winter peaks. Genome-wide association studies (GWAS) identified 4,024 susceptibility loci, potentially modulating immune-related gene expression to influence maternal antiviral immunity. These findings elucidate geographic and genetic influences on viral transmission, demonstrating the value of large-scale NIPT data in infectious disease surveillance. Our study provides critical evidence for establishing pregnancy infection warning systems and precision prevention strategies.

## Introduction

Published studies on the blood microbiome suggest the potential existence of an intrinsic core microbiota in the blood, not derived from contamination, which remains dormant in a healthy state but participates in physiological and immune regulation[1, 2]. Metagenomic next-generation sequencing (mNGS) of plasma mcfDNA holds clinical application prospects, enabling “unbiased” microbial detection through cell-free DNA (cfDNA) sequencing[3, 4]. Recent studies have detected viral genetic material across different populations, with 94 viral sequences identified in blood samples from 8,240 healthy donors[5] and 224 DNA viruses found in 108,349 prenatal specimens through NIPT analysis[6]. However, these studies had relatively small sample sizes (<100,000) and were cross-sectional , and the stability of viral prevalence in large populations has yet to be confirmed.

Furthermore, significant knowledge gaps persist regarding two core dimensions of infectious disease epidemiology—geographic heterogeneity and temporal dynamics—in the context of blood virome prevalence. Comparative studies have revealed significant regional variations in the distributions of viruses and microbes across China, including distinct divergence patterns of hepatitis B and C viruses (HBV-B/HBV-C) between the northern and southern regions[7], as well as marked differences in microbial diversity between Xinjiang and Beijing[8]. However, these studies were limited to spatial comparisons at single time points. Crucially, fundamental questions concerning the temporal dimension remain systematically unexplored. These include whether viral prevalence rates are influenced by seasonal cycles or interannual variations, and how environmental factors interact with host genetics. This limited understanding also hinders the development of effective infectious disease surveillance and early warning models.

Growing evidence indicates that complex genetic regulatory networks govern virus-host interactions[9]. Polymorphisms at specific loci can significantly alter individual infection risk by influencing viral receptor expression, immune response intensity, or the efficiency of viral genome integration[10, 11]. Human herpesvirus 6 (HHV6) chromosomal integration is strongly associated with the MLC1-MOV10L1 locus in infected individuals[12]. GWAS have revealed a missense variant in the ACR gene that is significantly linked to HHV6 infection, with additional suggestive signals near SHANK3 and RABL2B[7]. Furthermore, the rs11686168 variant in CDC42EP3 approaches statistical significance concerning human herpesvirus 5 (HHV5) reactivation risk post-transplantation[13]. Despite progress, limitations persist, including small sample sizes and narrow viral coverage. Crucially, genetic susceptibility loci for viral infections and their impact mechanisms on pregnancy outcomes remain unexplored in pregnant women, an immunologically distinct group. This research gap impedes the development of precision prevention strategies for prenatal infections.

To address these gaps, we analyzed sequencing data from 1,023,651 pregnant women who underwent NIPT at designated prenatal screening centers across Hebei Province, China, between 2019 and 2022. By aligning non-human cfDNA fragments against viral reference genomes, we constructed a virome map of the maternal blood in this large natural cohort. We revealed the epidemiological landscape across geographic and temporal dimensions. Furthermore, we investigated associations between viral infections and maternal baseline characteristics, explored patterns of viral co-infection, and identified genetic loci potentially linked to viral susceptibility through GWAS analysis. These findings offer novel insights for preventing viral infections during pregnancy.

## Results

### A population of one million characterized by diverse terrains and ethnic groups

We recruited a total of 1,023,651 pregnant women, approximately 90% of whom were from various prefecture-level cities in Hebei Province (Fig 1a). Hebei Province, which boasts the most comprehensive topography in China, encompassing plains, plateaus, mountains, hills, deserts, and coastal areas, had its pregnant women participants sourced from all topographical regions. The age of the pregnant women ranged from 20 to 45 years, with an average of 29.1 years, and about 11% were advanced maternal age (≥ 35 years). The gestational age at sampling was between 12 and 23 weeks, averaging 16.65 weeks. The participants represented 46 ethnic groups, with Han Chinese accounting for 93.8%, followed by Manchu, Hui, and Mongol at 4.21%, 0.84%, and 0.5%, respectively. The proportions of other ethnic minorities were all below 0.5%, consistent with the distribution characteristics of China’s 56 ethnic groups (Fig 1b).

**Fig 1.**
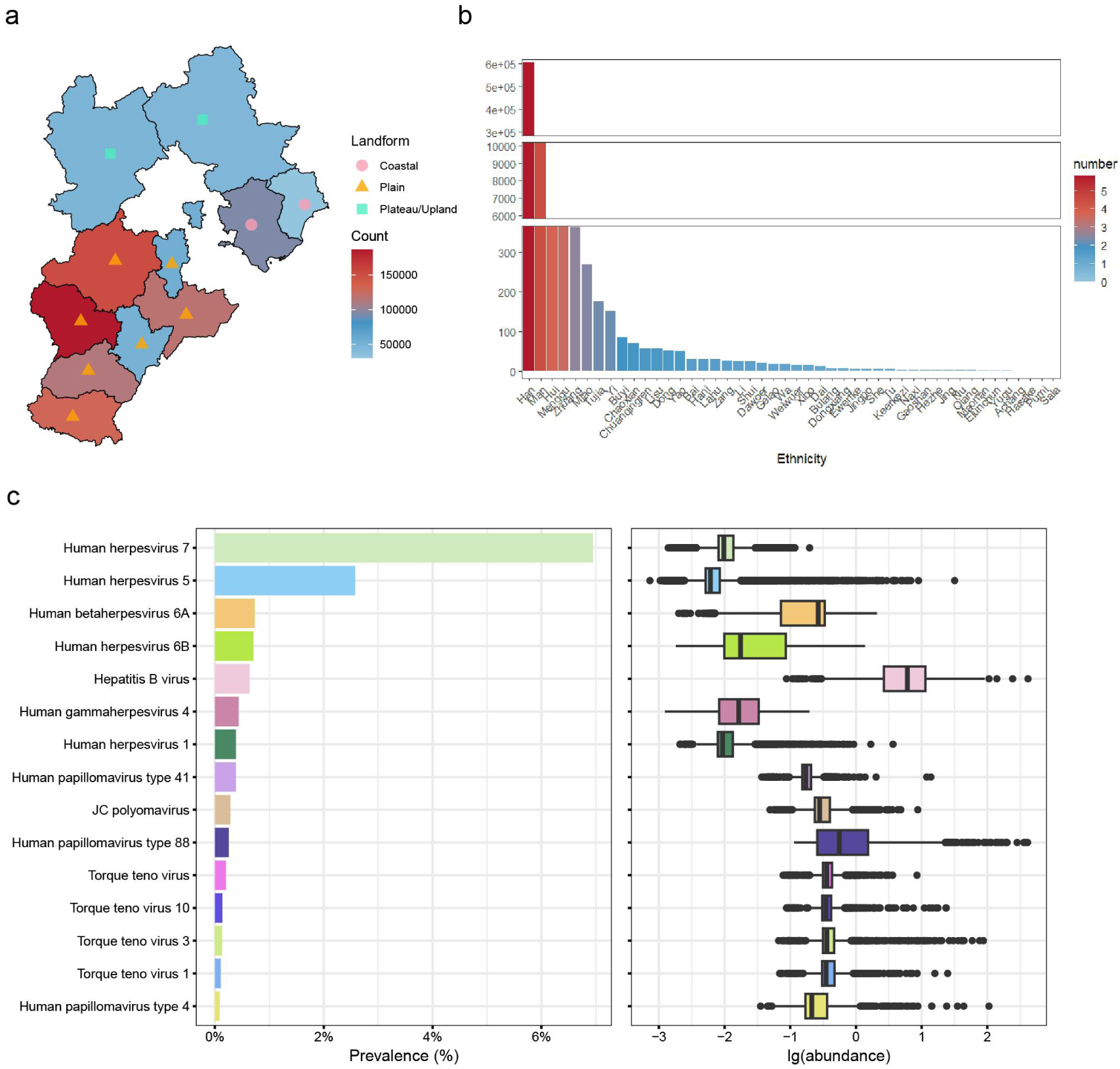
Demographic attributes and viral molecular prevalence within the investigated cohort. **a** Dissemination of the sample population across cities in Hebei Province. **b** Composition of ethnic groups and sample quantities. The illustration depicts the anglicized denominations of the ethnic minorities, with diverse symbols signifying varied geographical traits, and hue saturation correspondingly indicating the sample magnitude. **c** Hierarchical ranking of viral prevalence and relative abundance across millions of NIPT subjects. The left panel ranks the prevalence of various viruses in the blood from highest to lowest. The right panel shows the relative abundance of each virus in the blood, with each dot representing the viral abundance of an individual.

### The blood virome reveals high prevalence but low abundance of herpesviruses versus high abundance of HBV

Fig 1c illustrates the distribution of the top 15 most frequent viral pathogens and their respective relative abundances. Notably, human herpesvirus 7 (HHV7) constitutes 6.95% of the virome map, followed by HHV5 at 2.58%, HHV6 at 0.73%, and HBV at 0.64%, which collectively exhibit the highest detection frequencies. Detection of multiple types of torque teno virus (TTV) suggests a modest prevalence of TTV within the peripheral blood samples. HBV demonstrates the highest mean abundance, followed by JC polyomavirus and diverse TTV types. It is pertinent to observe that although several human herpesviruses are frequently detected, their mean abundances remain comparatively negligible.

### Gradual decline in viral prevalence across plains, plateaus, and coastal regions

Hebei Province has 11 prefecture-level cities, categorized by topography into 7 plain, 2 plateau, and 2 coastal areas. As shown in Fig 2a, HHV6, human papillomavirus (HPV), and Parvovirus B19 prevalence declines progressively from plains to plateau/upland, then coastal areas. TTV is more prevalent in the plateau/upland regions. HBV and HHV7 are more common in plains, with comparable but lower rates in plateau and coastal areas. Human herpesvirus 1/2 (HHV1/2) is relatively evenly distributed except for higher prevalence in plateaus (S1 Fig). Viral abundance across cities (Figs 2b, 2c) mirrors prevalence, with plains cities having higher levels. HBV and Parvovirus B19 show high abundance across all cities.

**Fig 2.**
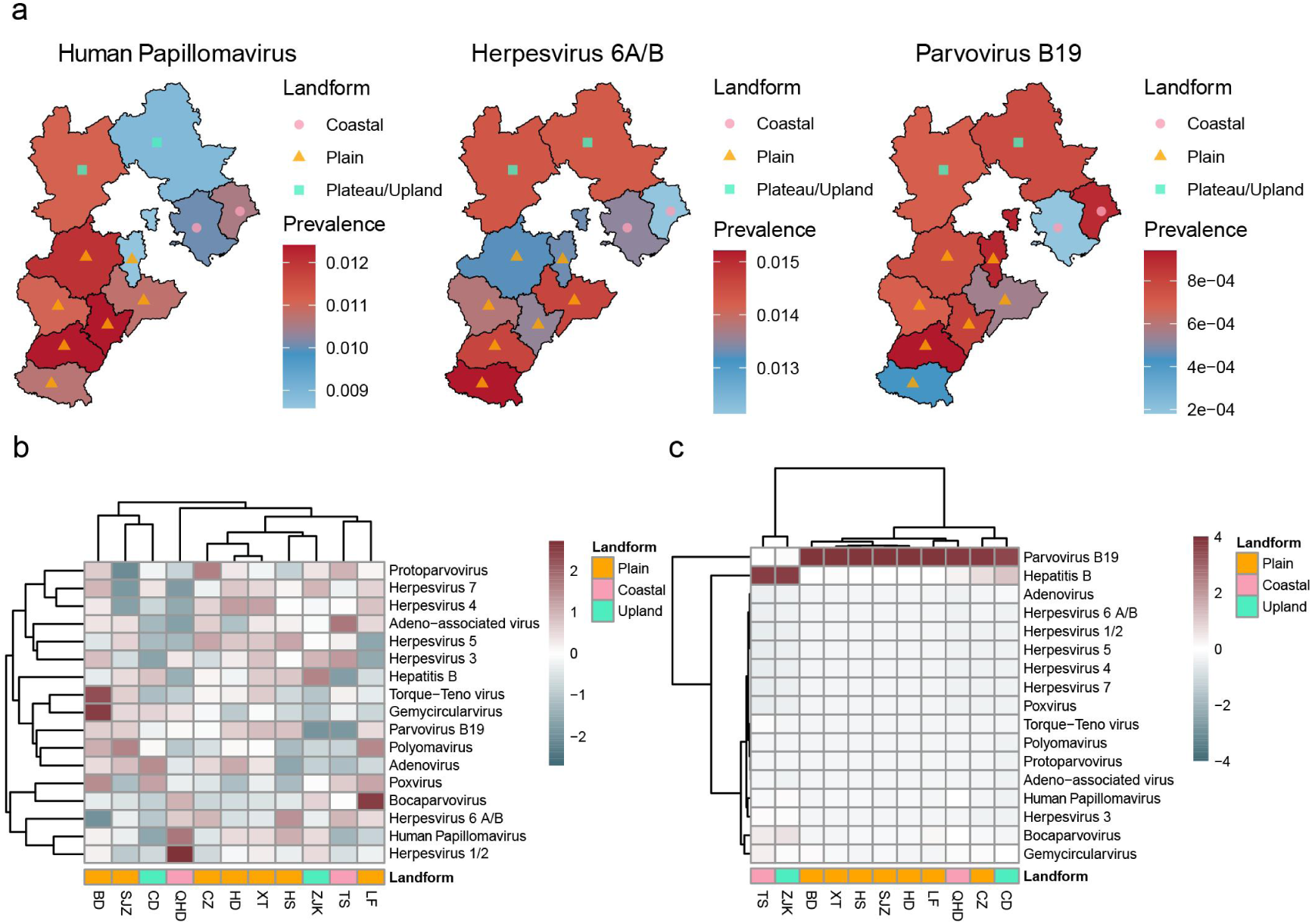
The epidemic characteristics of various viruses in terms of geographical distribution. **a** Heatmap of virus prevalence in cities of Hebei Province; triangles represent cities located in the plains; squares represent cities located in the plateau and upland regions; circles represent cities located in the coastal areas. **b** Heatmap of average viral abundance in cities of Hebei Province; each column represents the average viral abundance in each city. The more red the color, the higher the average viral abundance in that city. **c** Heatmap of average viral abundance in cities of Hebei Province; each row represents the average viral abundance of a particular virus across different cities. The more red the color, the higher the average viral abundance of that virus in those cities. SJZ, Shijiazhuang; BD, Baoding; CZ, Cangzhou; HD, Handan; XT, Xingtai; HS, Hengshui; LF, Langfang; TS, Tangshan; QHD, Qinhuangdao; CD, Chengde; ZJK, Zhangjiakou.

Moreover, an ethnic-specific analysis of viral prevalence was conducted, as detailed in S2 Fig. Except for certain ethnic cohorts, such as the Naxi and Xibo, in which no viral infections were detected in the blood samples, the Manchu ethnic group demonstrated significantly elevated infection rates for a diverse array of viruses relative to other ethnic groups. HHV5 and HHV7 emerged as the predominant viral infections consistently across all ethnicities, corroborating the observations from the expansive virome map.

### Seasonal pattern of viral prevalence: gradual increase from summer to autumn and winter

From 2019 to 2022, the number of pregnant women included in our study was 176,404, 404,504, 219,357, and 223,386, respectively. Following the omission of data for the period from January to May 2021, due to its incomplete nature, we graphically represented the temporal fluctuations in viral prevalence (Fig 3a, S3 Fig). Analyzed results indicate that the prevalence of HHV7, HHV5, HHV4, and TTV increased in 2021 and 2022. Specifically, HHV7 and HHV5 showed higher prevalence each year in November; TTV exhibited higher prevalence in November and February; and human herpesvirus 4 (HHV4) showed a significant increase in prevalence after March 2022. The prevalence of other viruses fluctuated minimally over time.

**Fig 3.**
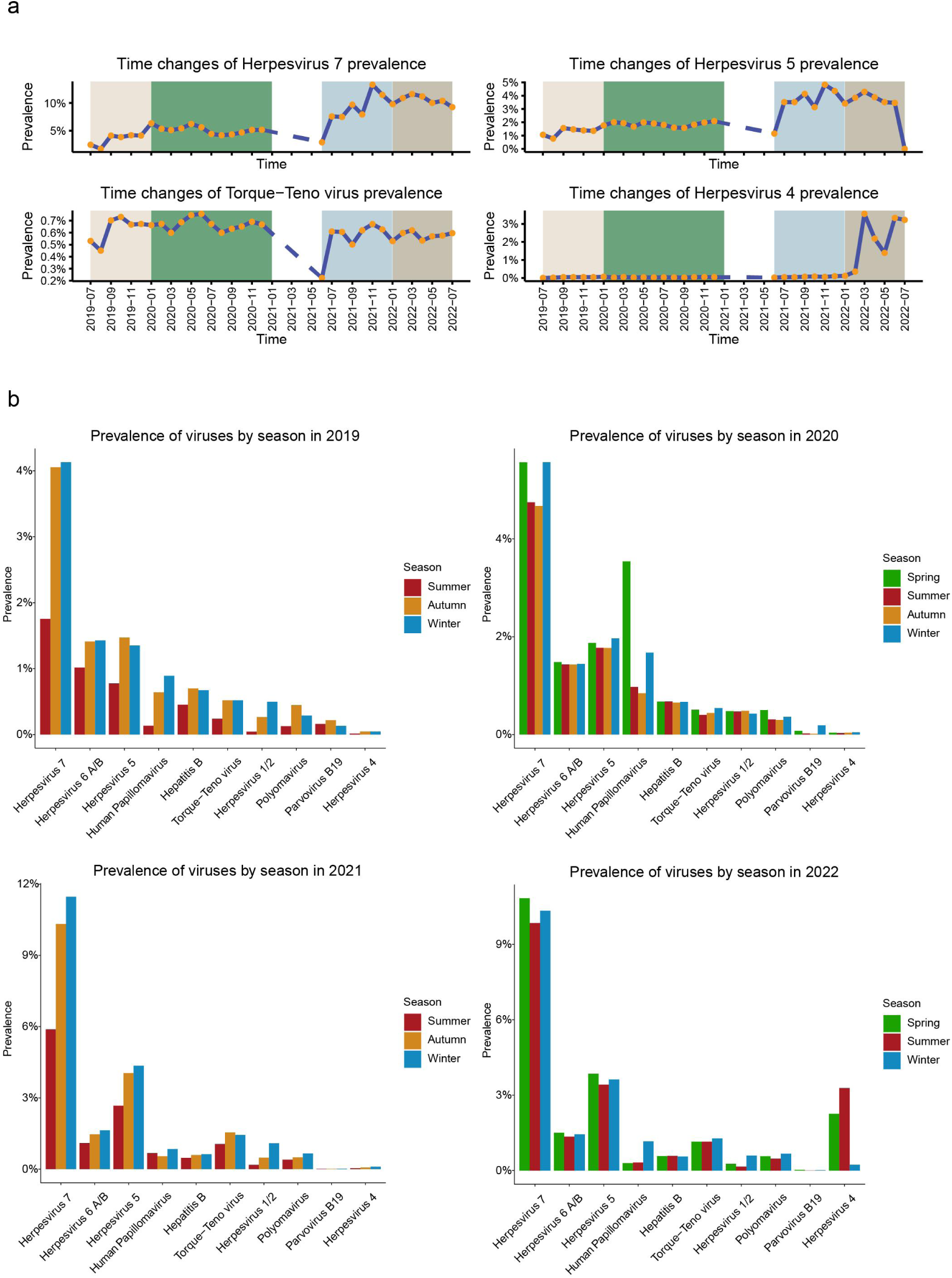
The epidemic characteristics of various viruses over time. **a** Comparison of viral prevalence across different years. **b** Comparison of viral prevalence across different seasons in each year.

From a seasonal perspective, between 2019 and 2022, the prevalence of most viruses were higher in spring, autumn, and winter than in summer, and each year exhibited distinct seasonal patterns (Fig 3b). In 2019, viruses such as HHV7, HHV6, HPV, and HHV1/2 showed a seasonal increase from summer to autumn to winter. In 2020, HHV7 and HHV5 were more prevalent in spring and winter, while HHV6, HBV, and HHV1/2 remained stable. The trend continued in 2021, with most viruses exhibiting higher prevalence from summer to autumn to winter. In 2022, except for HHV4, other viruses were more common in winter and spring than in summer.

### Correlation between virus infection rate and maternal health status during pregnancy

Logistic regression analysis was employed to investigate the associations between maternal age, height, weight, body mass index (BMI), gestational week (GWK) at NIPT sampling, and viral infections, as depicted in S4a Fig and S1 Table. The findings indicated that the likelihood of HHV7, HHV5, polyomavirus, and HBV infections diminished with advancing maternal age, whereas the probability of HHV6 infection augmented with increasing age. Maternal height was positively correlated with the risk of HHV7 infection and inversely associated with HHV5 infection risk. The likelihood of HHV7 infection negatively correlated with maternal weight, whereas HHV6 infection risk was positively associated with weight. Furthermore, the probability of polyomavirus infection was found to escalate with advancing GWK.

We also compared the changes in viral prevalence across different gestational age groups (S5 Fig). We found that whether it was appropriate gestational age (20-34 years) or advanced gestational age (≥ 35 years), the viruses with the highest prevalence were HHV7, HHV6, CMV, and HBV, showing almost no difference from the overall results. However, HHV7, CMV, HBV, polyomavirus, and TTV had higher prevalence rates in the appropriate gestational age group, while HHV6 and HHV4 had higher prevalence rates in the advanced gestational age group, which is consistent with the results of the logistic regression analysis.

### Co-infection patterns among different viruses

Among the million pregnant women samples, 386,046 exhibited multiple viral co-infections, with a broad distribution across periods and regions (S4b Fig). Analysis of co-infection patterns (S4c Fig) revealed 17 viral genera with co-infection frequencies exceeding 50 instances. HPV, HHV7, HHV5, and polyomavirus showed high co-infection propensity, with HPV having the highest likelihood. HHV7 and HHV5 displayed the widest range of co-infection frequencies and diversity, and viruses with higher co-infection propensity tended to co-infect each other more frequently.

### Human leucocyte antigen (HLA) family as susceptibility factors for multiple viral infections

GWAS identified 88, 2198, 1005, 420, 306, and 7 significant loci associated with susceptibility to HHV6B, HHV7, HBV, TTV, HHV5, and HPV, respectively (Fig 4a, S2-3 Tables). For HHV6B, 87 genome-wide significant loci (p < 5×10^−8^) were found on chromosome 22, consistent with previous reports[12], primarily enriched in MLC1 (21 significant loci) and MOV10L1 (27 significant loci). In addition to identifying the previously recognized viral genetic susceptibility loci, our study uncovered multiple novel variants potentially linked to viral infections.

**Fig 4.**
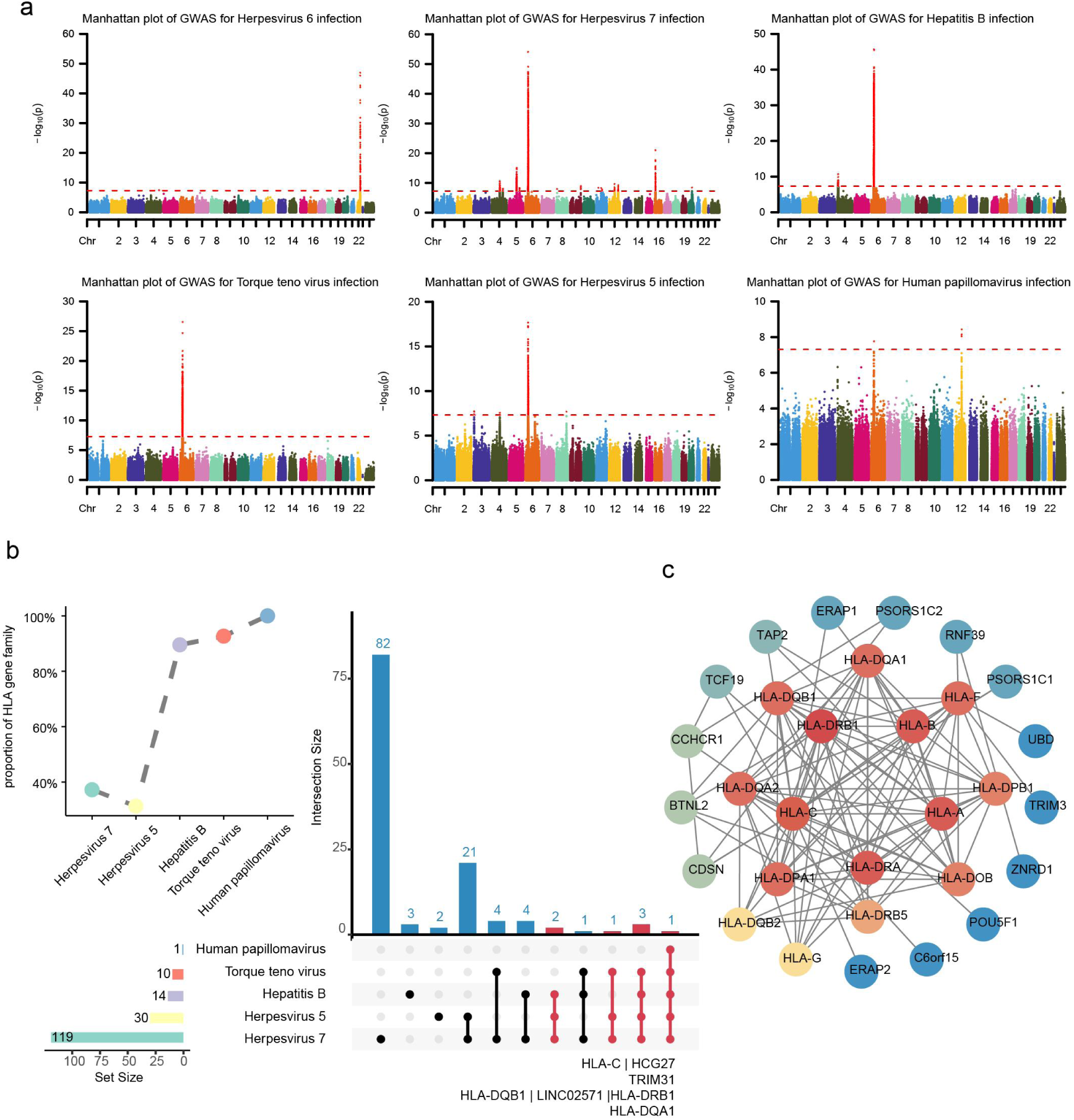
Genetic susceptibility loci, gene features of multiple viruses, and networks of HHV7, HHV5 & HBV. **a** Manhattan plots for the significant susceptibility loci of HHV6B, HHV7, HBV, TTV, HHV5, and HPV, respectively. **b** Upset plot showing overlap of susceptibility genes across five viruses, and proportion of HLA genes among susceptibility genes for each virus. **c** The interaction network of three viral proteins predicted by the STRING database (threshold: 0.7).

For instance, we have newly identified 2,198 GWAS significant loci associated with HHV7 infection (Supplementary Tables 2-3). Notably, the variants on chromosome 4 were primarily enriched in the NFKB1 and MANBA regions. NFKB1 encodes NF-κB, a critical regulator of early immune responses to viruses through alternative splicing and protein processing[14], while MANBA may modulate the viral replication environment via the lysosomal metabolic pathway[15]. Furthermore, the association sites on chromosome 5 were predominantly located in the ERAP1/ERAP2 region. These genes encode aminopeptidases that play a crucial role in trimming antigenic peptides, thereby participating in the antigen presentation process of HLA class I molecules. Dysfunction of these genes has been associated with pregnancy complications, particularly preeclampsia[16, 17]. The highest density of association loci was observed on chromosome 6 within the major histocompatibility complex (MHC) region, which encompasses multiple HLA complex subtype genes, including HLA-DRB1 and HLA-DQA1.

It is noteworthy that significant peaks for five viruses, including HHV7 and HBV, were detected on chromosome 6. Some of these significant loci are shared, primarily distributed in immune-related genes such as HLA-C, TRIM31, HLA-DQB1, and HLA-DQA1 (Fig 4b). The HLA family genes account for more than 30% of the virus-associated genes across all viruses (Fig 4b), among which HLA-DQA1 is significantly associated with susceptibility to these viruses.

### Host genetic susceptibility loci modulate immune responses via HLA family genes

Expression quantitative trait loci (eQTL) associated with 22 genes linked to HHV7, HHV5, and HBV infections were identified in the GTEx database (Supplementary Table 4). These eQTL loci potentially modulate gene expression in tissues such as skin and heart. Using genotype data from 3202 individuals in the International 1000 Genomes Project, LD between these eQTL loci and GWAS-significant loci was calculated. Results indicated high LD (r^2^ > 0.8) for 7, 1, and 20 eQTL loci in the HLA-DQB1, HLA-DRB1, and HLA-C regions, respectively, with their corresponding GWAS loci.

Additionally, 124 genes were associated with GWAS-significant loci for the three viruses. A Protein-protein interaction (PPI) network was constructed via the STRING database. Fig 4c depicts 30 nodes with a degree score >0, revealing a dense cluster formed by 15 HLA genes (e.g., HLA-DQB1, HLA-DRB1, HLA-C), two transcription factors (POU5F1 and TCF19), and 13 other GWAS-associated genes. Further, gene expression data from the GSE83148 dataset were collected, organized, and used to construct a co-expression network following RMA normalization. Of the 124 GWAS-associated genes analyzed, 78 were successfully mapped onto the co-expression network. Within this subset, 57 genes formed a fully connected sub-network, comprising 379 co-expression relationships. Notably, HLA family genes—including HLA-C, HLA-B, HLA-H, and HLA-DRB—along with other GWAS-associated genes, formed a densely connected and stable co-expression cluster (Fig 5).

**Fig 5.**
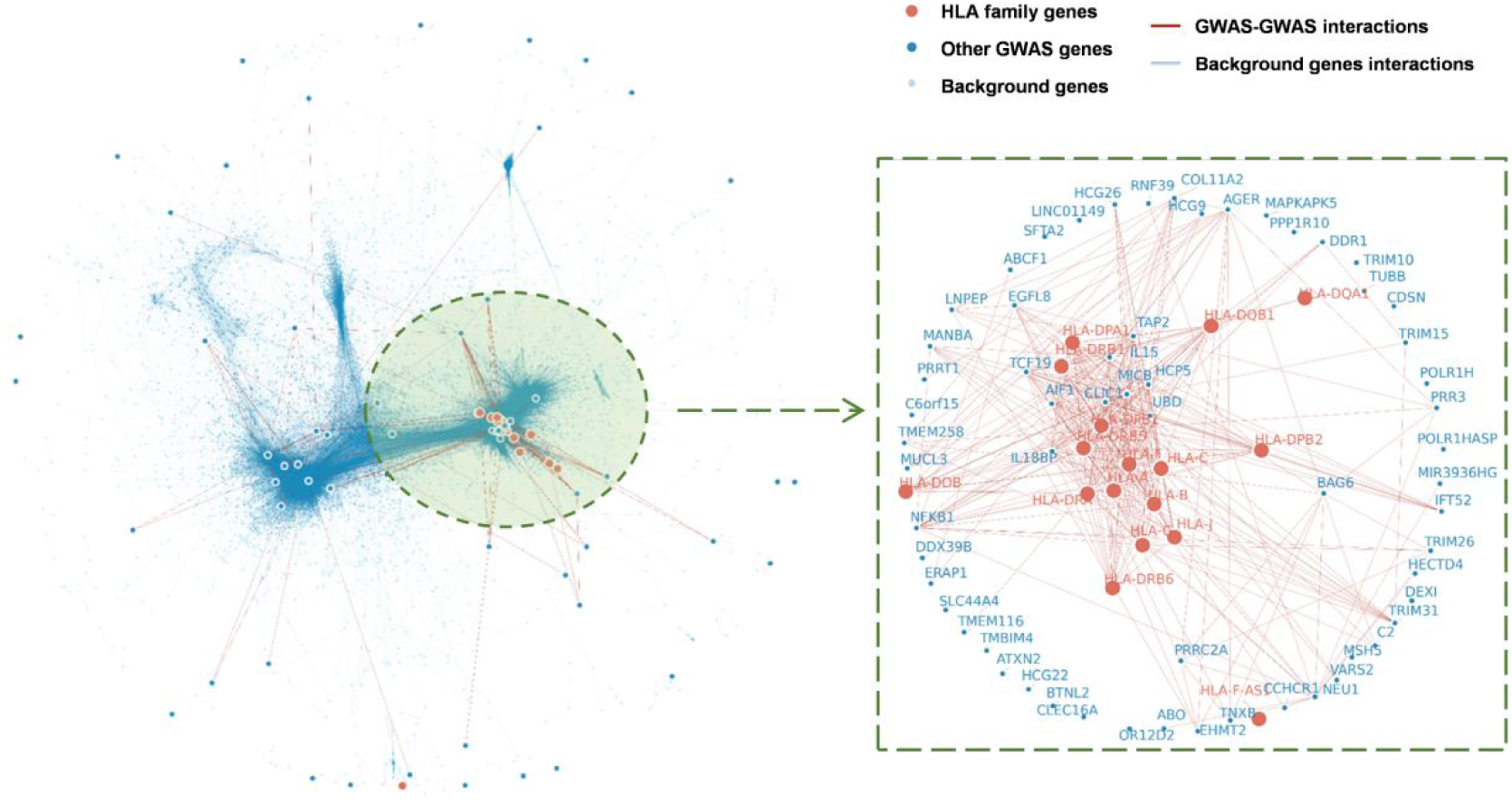
Weighted gene co-expression network analysis of GSE83148. The left figure shows the WGCNA co-expression network (GSE83148). GWAS genes are represented by large red (HLA family genes) or blue (other genes) nodes. Red lines represent interactions between GWAS genes. Small light blue nodes and lines represent background genes and their interactions. The right figure shows a subnetwork containing only GWAS genes; the symbols and colors are the same as the left figure. The visualization reveals a highly interconnected subnetwork, with HLA genes and other GWAS-associated genes occupying a central position, forming a densely connected module.

The 124 GWAS-associated genes that exhibit PPI and co-expression relationships with these HLA family genes were subjected to Gene ontology (GO) enrichment analysis. The results revealed that these genes are involved in immune-related functions, particularly antigen presentation. Within the biological process category, the most significantly enriched terms were related to antigen processing and presentation, especially “antigen processing and presentation of peptide antigen” (S6 Fig). In the cellular component category, the significantly enriched terms primarily involved membrane structures and transport vesicles (S6 Fig). In the molecular function category, several terms associated with antigen binding and peptide binding also showed significant enrichment (S6 Fig).

## Discussion

Our study is based on the NIPT data of one million pregnant women in Hebei Province from 2019 to 2022, constructing the largest-scale blood virome map of a pregnant population globally. Combining the diverse topographic features of Hebei Province and the characteristics of 46 ethnic groups residing there, we analyzed the impact of geographic and ethnic factors on viral infections. The research also explored the associations between infections and clinical data, co-infection patterns of viruses, as well as genetic susceptibility loci and related mechanisms. These findings enhance our understanding of the viral composition and epidemic patterns within this vast pregnant population.

The virome map has for the first time revealed the distribution patterns of viruses in the blood of one million pregnant women in Hebei Province, with a higher prevalence of human herpesviruses such as HHV7 and HHV5—consistent with European studies on the general healthy population[6], suggesting that the viral composition in pregnant women’s blood may be similar to that of the normal population[5]. Therefore, the large-scale blood virome map of pregnant women reflects the natural viral infection patterns in the population. Compared with Chinese studies on viral infections in the blood of pregnant women, our findings differ, showing a higher prevalence of HHV7/HHV5 and a lower prevalence of HBV. China is a high-burden country for HBV, with a higher prevalence of HBV than Europe[18]. Recent government prevention strategies have curbed the spread of HBV, demonstrating that effective policies can significantly reduce the infection rates of viruses such as HBV[18, 19].

Geographically, the diversity and abundance of viruses in plain regions are higher than in other terrains. The prevalence of HHV7, HHV6, and HPV decreases sequentially from plains to highland mountainous areas and then to coastal regions. The convenient transportation and dense population in plain areas facilitate virus transmission[20], while the isolation and lower population density in highland regions slow down virus spread[21, 22]. Despite the dense population in Hebei’s coastal area, stronger ultraviolet radiation and faster air circulation contribute to a lower viral prevalence. While metagenomic studies have confirmed that geography, climate, and spatial distance affect the diversity and variation of bacterial communities[23, 24], research into the geographical distribution of viruses remains insufficient. Our analysis of virus prevalence in different landform environments reveals that the distribution of viral infections in geographical space exhibits distinct localized characteristics.

Temporal analysis shows increased viral prevalence in 2021-2022, coinciding with Hebei’s peak coronavirus disease 2019 period. The Severe acute respiratory syndrome coronavirus 2 may impair innate immunity[25], while prolonged preventive measures (e.g., masks, disinfection, distancing) likely reduced pathogen exposure, lowering antibody levels and increasing susceptibility post-restriction easing[26]. Further research is needed to confirm these mechanisms and guide control strategies. In addition, viruses exhibit clear seasonality, peaking in spring/winter and declining in summer. Colder temperatures enhance viral survival and transmission[27], while increased indoor gatherings facilitate spread[28, 29]. These insights underscore the value of large-scale, time-resolved mcfDNA data for advancing epidemic surveillance and early warning systems.

Our investigation into viral distribution in pregnant blood samples revealed a notable co-occurrence of HPV, HHV7, HHV5, and polyomavirus. This phenomenon of co-infection appears to be intricately linked to their replication strategies, host adaptation mechanisms, and inter-viral interactions. Specifically, HPV employs a layered replication strategy, maintaining latent, low-copy plasmids in basal cells to facilitate persistent infection, thereby creating an “immunological shield” that may benefit viruses such as HHV7[30]. Concurrently, HHV7 exhibits a dual infection mechanism, achieving latency in CD4+ lymphocytes while actively replicating in salivary glands[31], leading to persistent viremia that can support the infection of other viruses like HHV5[32]. Moreover, BK polyomavirus potentially augments the replication of viruses, including HHV5, through shared host transcription factors or microRNA networks[33–35]. These findings underscore the complexity of preventing viral co-infections in large populations and highlight the urgent need for further foundational research into the mechanisms governing these viral interactions.

Our comprehensive GWAS analysis of infections, encompassing HHV6B, HHV7, HBV, TTV, HHV5, and HPV, identified 4,024 genetic susceptibility loci, with a notable 30% situated within the HLA gene family region. Through colocalization analysis, we uncovered robust associations between skin tissue eQTLs of genes such as HLA-DQB1, HLA-DRB1, and HLA-C and their corresponding GWAS loci. The skin, serving as the primary immune barrier against pathogen invasion[36], leverages HLA family genes to encode cell-surface antigen-presenting molecules, thereby initiating immune responses[33]. These insights suggest that the newly identified viral susceptibility loci may play a pivotal role in modulating pregnant women’s antiviral immunity by influencing HLA gene expression.

PPI network analysis unveiled that HLA family genes engage in close interactions with other GWAS-associated genes. Subsequent co-expression network analysis indicates that HLA genes may intricately modulate immune responses through cooperative mechanisms with these genes. For instance, ERAP1 and ERAP2 are involved in the processing of antigen peptides, thereby influencing HLA class I antigen presentation[37]. Additionally, the transcription factor TCF19 plays a regulatory role in HLA gene expression, impacting downstream immune cascades[38]. In summary, our findings pinpoint novel potential targets for precise viral control, warranting further in-depth investigation.

Our study demonstrates the potential value of mcfDNA in uncovering spatiotemporal patterns of viral infection and epidemiological surveillance, while acknowledging several limitations. First, the absence of paired serological validation precludes accurate assessment of NIPT’s sensitivity and specificity for viral detection. Second, the mechanisms underlying geographic disparities in viral infection rates remain to be elucidated due to insufficient data on individual lifestyles and environmental factors. Finally, the lack of information on pregnancy outcomes and gestational complications necessitates further investigation into the effects of viral infections on pregnancy-related disorders in large-scale populations.

In the future, we plan to systematically collect clinical diagnostic information, examination and test results, and demographic data during pregnancy. This comprehensive data collection will enable us to more accurately delineate the spatiotemporal patterns of viral infections and their impact on maternal health. Additionally, we will further develop and optimize mcfDNA detection technology to enhance the accuracy of pathogen identification, thereby providing a more robust surveillance tool for clinical practice and public health initiatives.

## Materials and methods

### Sample information

In adherence to the Regulations of the People’s Republic of China on the Management of Human Genetic Resources, the Health Commission of Hebei Province has furnished a collection of 1,023,651 anonymized samples, procured from 2019 to 2022. This dataset consists of raw FASTQ files derived from NIPT, complemented by corresponding sample metadata. The clinical parameters encompass maternal age, ethnicity, registered place of residence, location of the screening institution, height, weight, BMI, gestational age, and the date of sample collection. BMI was computed using the conventional formula: “Weight (kg) divided by height squared (m²).” In cases where participants had multiple NIPT records, data from the most recent examination were exclusively utilized.

### NIPT sample collection, sequencing, and data alignment

For NIPT, 5 mL of peripheral blood was collected from each participant into EDTA-anticoagulant tubes. Samples underwent immediate two-step centrifugation: first at 1,600 × g for 10 minutes at 4℃ to separate plasma, followed by transfer of the supernatant to new tubes for high-speed centrifugation at 16,000 × g for 10 minutes to remove residual cellular debris[39]. Subsequently, 200 μL of clarified plasma was aliquoted from each sample for cfDNA extraction. Library construction was performed on the MGISEQ-2000 platform using DNA nanoball technology with probe-anchored synthesis. This process included library preparation, quality assessment (encompassing fragment size distribution analysis and concentration quantification), and library pooling. Final high-throughput sequencing employed a 35-bp single-end read configuration[40].

The raw sequencing data were subjected to quality control via SOAPnuke (version 1.5.6), wherein reads were excluded if they met either of the following criteria: more than 10% of bases possessed quality scores (Q) below 5, or contained ambiguous bases (N). After rigorous quality control measures, an average of 6.86 × 10⁶ reads per sample (corresponding to 2.40 × 10⁸ bases) were deemed qualified, resulting in a genomic coverage depth of 0.08-fold. These quality-controlled reads were then aligned to the GRCh38 reference genome, excluding alternative loci, utilizing BWA-MEM (version 0.7.17) with the parameters -T 28 -M. After alignment, read sorting and duplicate marking were executed with Sambamba (version 0.8.0). Variant detection was optimized by employing the GATK toolkit (version 4.1.2.0), encompassing: (1) local realignment of reads flanking known insertion/deletion (Indel) sites through the Indel Realignment module, and (2) systematic recalibration of base quality scores through the Base Quality Score Recalibration module[41].

### Single nucleotide variants (SNV) calling

In light of the propensity for non-uniqueness in alignments derived from single-end short-read sequencing data, this investigation scrupulously adhered to the analytical protocols standardized by the Global Alliance for Genomic Health Benchmarking Team, the Genome in a Bottle Consortium, and the Telomere-to-Telomere Consortium. The analysis was specifically calibrated for 35-bp reads, utilizing the mappable regions BED file derived from the GRCh38 reference genome, which encompasses 2,048,777,209 base pairs and was constructed using the GEM software library[42]. The resultant mean coverage depth comprised 7.53% of the genome, corresponding to 11.21% of the mappable regions. The initial SNV calling, executed with Base Var software on the alignment outcomes, produced 68,873,369 raw SNV loci, among which 44,907,889 were situated within mappable regions. Following iterative filtering with a quality score threshold (QUAL<80), a high-confidence dataset comprising 39,710,966 SNVs was finally compiled.

### The million-pregnant-women blood virome map construction

For each NIPT sample mapped to the GRCh38 reference genome, sequences of non-human origin are excised utilizing Samtools (version 1.12). Subsequently, these sequences are aligned with 224 human-hosted DNA viral genomes[6], using BWA (version 0.7.17), retaining only those reads uniquely mapped to viral genomes. The prevalence and relative abundance of viruses are computed following the strategy proposed by Guo[7] and Liu[12]. During the prevalence calculation, a sample is designated as a carrier of a virus if at least two sequencing reads specifically map to a particular viral reference genome. The formula for calculating relative abundance is as follows:

Abundance = 2 × [(Number of reads mapped to viral genome / Viral genome size) / (Number of reads mapped to hum

### Logistic regression analysis

Maternal age, BMI, weight, height, and gestational age were standardized using Z-scores. Logistic regression models were constructed using R’s stats package to analyze associations between these factors and carriage status of the 10 most prevalent viral genera, including HHV7 and HHV5. Viral carriage status (the dependent variable) was defined as binary (positive/negative) according to the criteria established in the million-pregnant-women blood virome map. The statistical significance of associations was assessed through regression coefficients and their hypothesis tests (p < 0.05 considered non-significant). All analyses were implemented using an in-house script.

### GWAS

GWAS for HHV6B, HHV7, HBV, TTV, HPV, and HHV5 were performed using PLINK2. The analysis employed a mixed linear model with viral carrier status (positive/negative) as the dependent variable and SNP genotype as the independent variable. Covariates included age, BMI, GWK at sampling, and the top ten principal components. Statistical significance for genome-wide associations was set at a P-value threshold of 5 × 10⁻⁸. The association results were visualized using the R package CMplot, generating Manhattan plots for genome-wide signal representation and Quantile-Quantile plots to assess population stratification and model fit quality.

### GO enrichment analysis

In GWAS, candidate susceptibility genes are identified as those closest to loci significantly associated with viral susceptibility (P < 5×10^−8^), within a 50-kilobase interval bordering these loci. Subsequent GO enrichment analysis was conducted using the enrichGO function from the R package clusterProfiler, with P-values corrected by the Benjamini-Hochberg method. The results were filtered to include entries with a P-value ≤ 0.05, an odds ratio ≥ 2, and gene set sizes ranging from 10 to 500.

### Colocalization analysis

For the 22 HLA susceptibility genes (including HLA-DQB1, HLA-DRB1, and HLA-C) implicated by GWAS, their eQTLs were retrieved from the GTEx database (version 10). Using PLINK 2.0 software, genotype data for these eQTL variants and the GWAS-significant variants associated with these genes were extracted from the 1000 Genomes Project Phase 3 data[43]. Subsequently, pairwise linkage disequilibrium (LD) levels were calculated between these variants. SNP pairs exhibiting an LD value (r²) > 0.8 were defined as having a strong colocalization relationship.

### PPI networks construction

The 124 associated genes identified by GWAS analysis for HHV7, HHV5, and HBV viruses were input into the ‘multiple proteins’ search module of STRING (v12.0), with the host species set to Homo sapiens and an interaction score threshold ≥0.7. After filtering out isolated nodes, the interaction data was exported in TSV format. Subsequently, network visualization analysis was performed using Cytoscape (v3.9.1) utilizing the built-in “Analyze Network” tool to calculate network topology parameters. Color gradients were mapped based on node Degree values, with red indicating high Degree values and blue indicating low Degree values. Finally, a hierarchical circular layout was employed to visualize the PPI network.

### Co-expression network construction

The GSE83148 dataset, obtained from the GEO database and sequenced on the GPL570 platform, was chosen for analysis. This dataset comprises 128 samples, including 122 HBV-infected individuals and 6 healthy controls[44]. Affymetrix CEL files underwent preprocessing using the RMA algorithm, executed via the “oligo” package, to perform background correction and normalization. Probe signals were then mapped to gene symbols using the “hgu133plus2.db” annotation package, with signals averaged accordingly. Quality control was conducted using the “limma” and “flashClust” packages, leading to the identification and removal of outlier samples. A signed co-expression network was subsequently constructed using the WGCNA package[45], with parameters set to a minimum module size of 30 genes and a merge height of 0.25. Edges with a topological overlap measure exceeding 0.001 were retained to form the network. The final co-expression network encompassed 16,503 genes linked by 7,908,425 co-expression relationships.

## Data Availability

All data produced in the present study are available upon reasonable request to the authors.
All data produced in the present work are contained in the manuscript.

## Author contributions

Conceptualization: Jianguo Zhang, Xiangdong Xu, Lijian Zhao, Jing Liu, Jianhong Zhao

Data analysis: Zhixu Qiu, Kaixuan Duan, Shaohui Qian, Shuo Li, Zhe Lin, Hankui Liu

Data Curation: Meng Wu, Sijie He Yanan Niu, Cuixin Qiang, Pu Qin, Zhirong Li, Jing Yang, Weigang Wang, Yunfang Wang, Defeng Tao, Chengbin Yan, Shuyang Gao,

Resources: Xiangdong Xu,Jing Liu, Jianhong Zhao

Writing and review: Zhixu Qiu, Kaixuan Duan, Shaohui Qian, Shuo Li, Linfeng Yang

Funding acquisition: Jianguo Zhang, Lijian Zhao All authors read and approved the final manuscript

## Ethies statement

The written informed consent was approved by the Institutional Review Board of BGl Genomicsand obtained from each participant. The study was approved by the Institutional Review Board of BGI Genomics and Hebei General Hospital. The genetic data collection and GWAS summary sharing was approved by the Human Genetic Resources Administration of China: [2023]CJ0271

## Acknowledgements

We thank the participants for their involvement in this study. This study was supported by S&T Program of Shijiazhuang(235790429H). We are grateful to Dr. Ruifu Yang (State Key Laboratory of Pathogen and Biosecurity, Academy of Military Medical Sciences) for his insightful comments regarding this work.

---

**S1 Fig.**
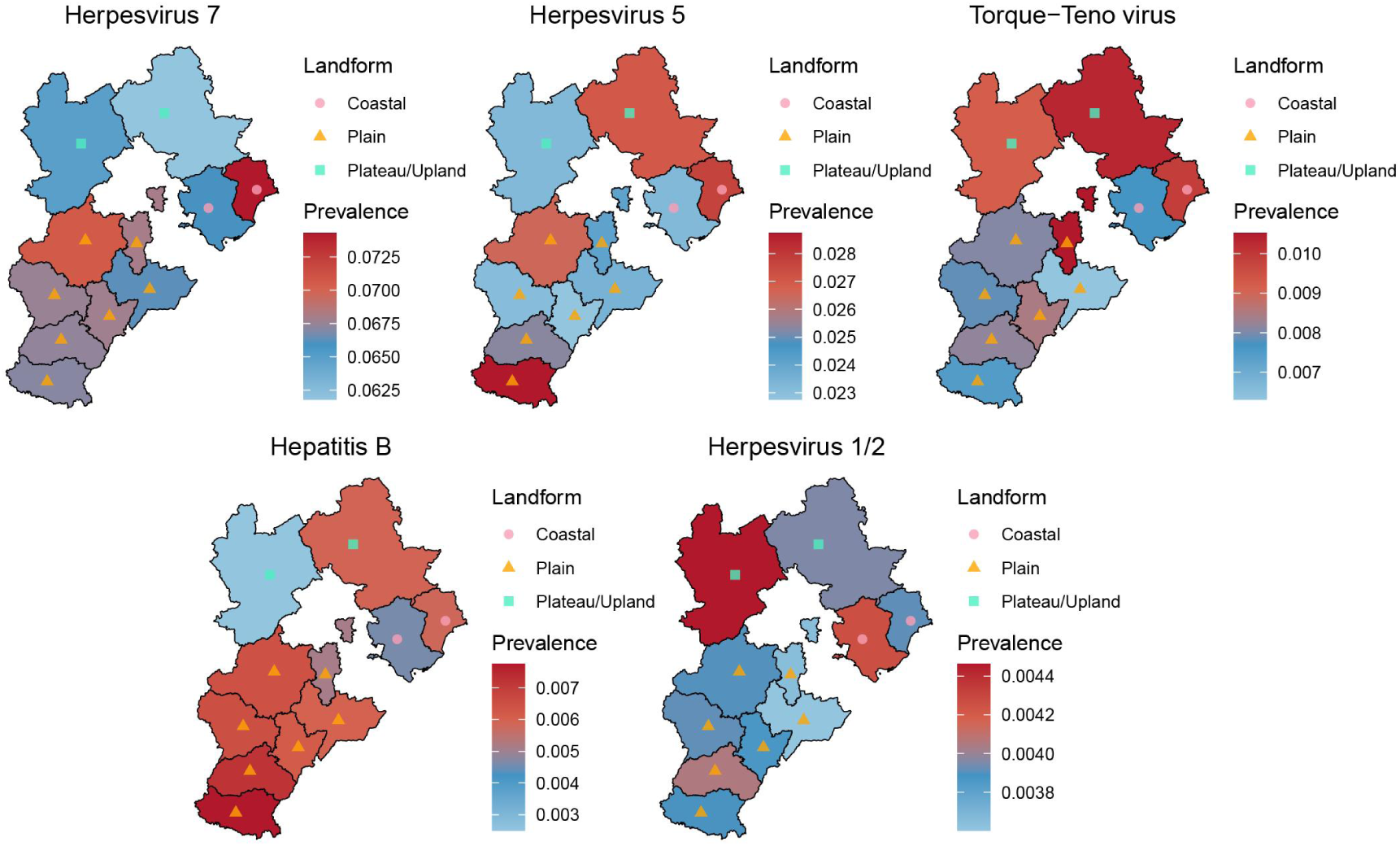
The heatmap of viral prevalence distribution across cities in Hebei Province. Triangles represent cities with plain terrain, squares represent cities with plateau and upland terrain, and circles represent coastal cities.

**S2 Fig.**
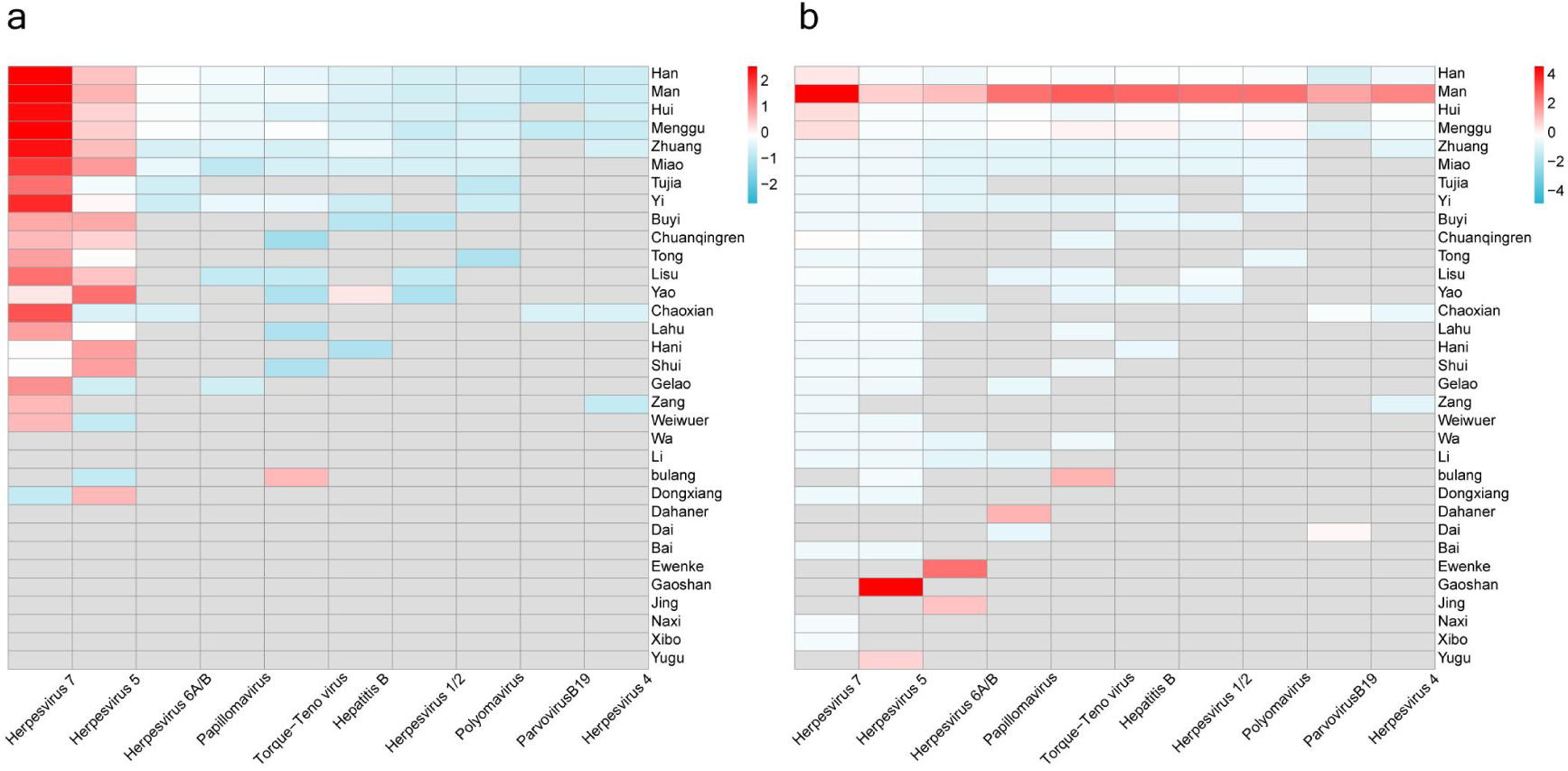
Heatmap of the distribution of infection numbers for each virus across different ethnic groups. **(a)** Each column represents the infection count of a specific virus across ethnic groups, with redder colors indicating higher infection rates. (**b)** Each row represents the infection count of viruses within a specific ethnic group, with redder colors indicating a higher viral infection rate in that ethnic group.

**S3 Fig.**
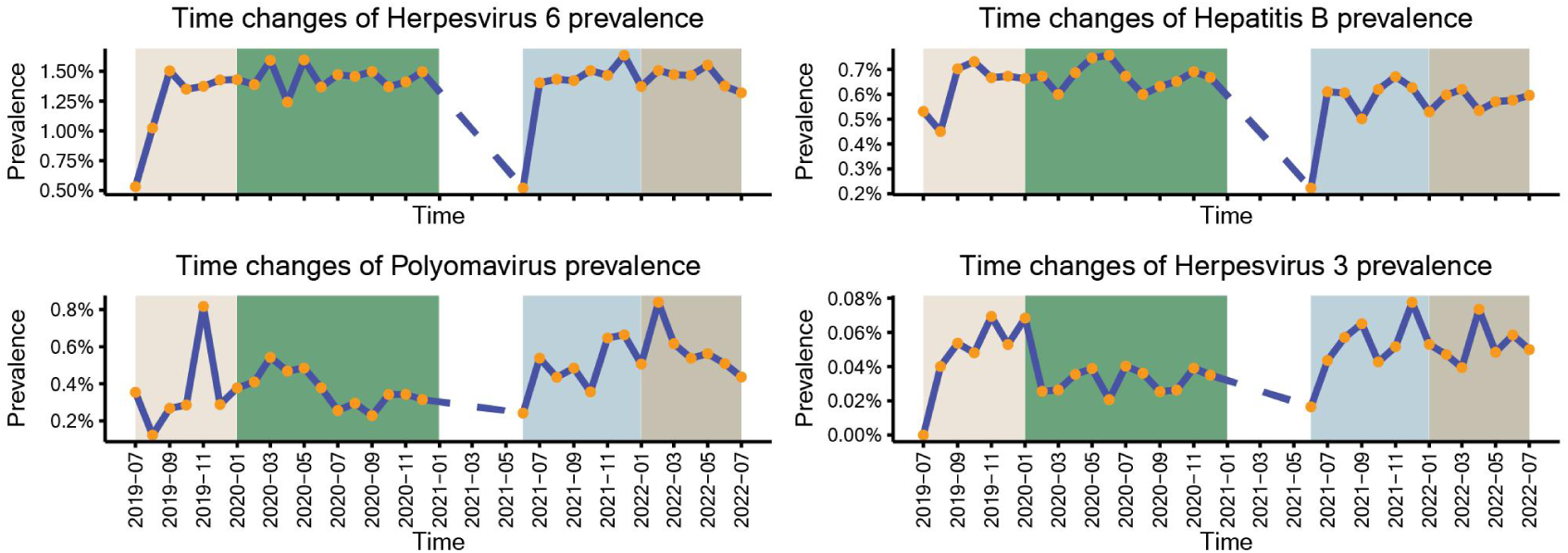
Comparison of viral prevalence across different years.

**S4 Fig.**
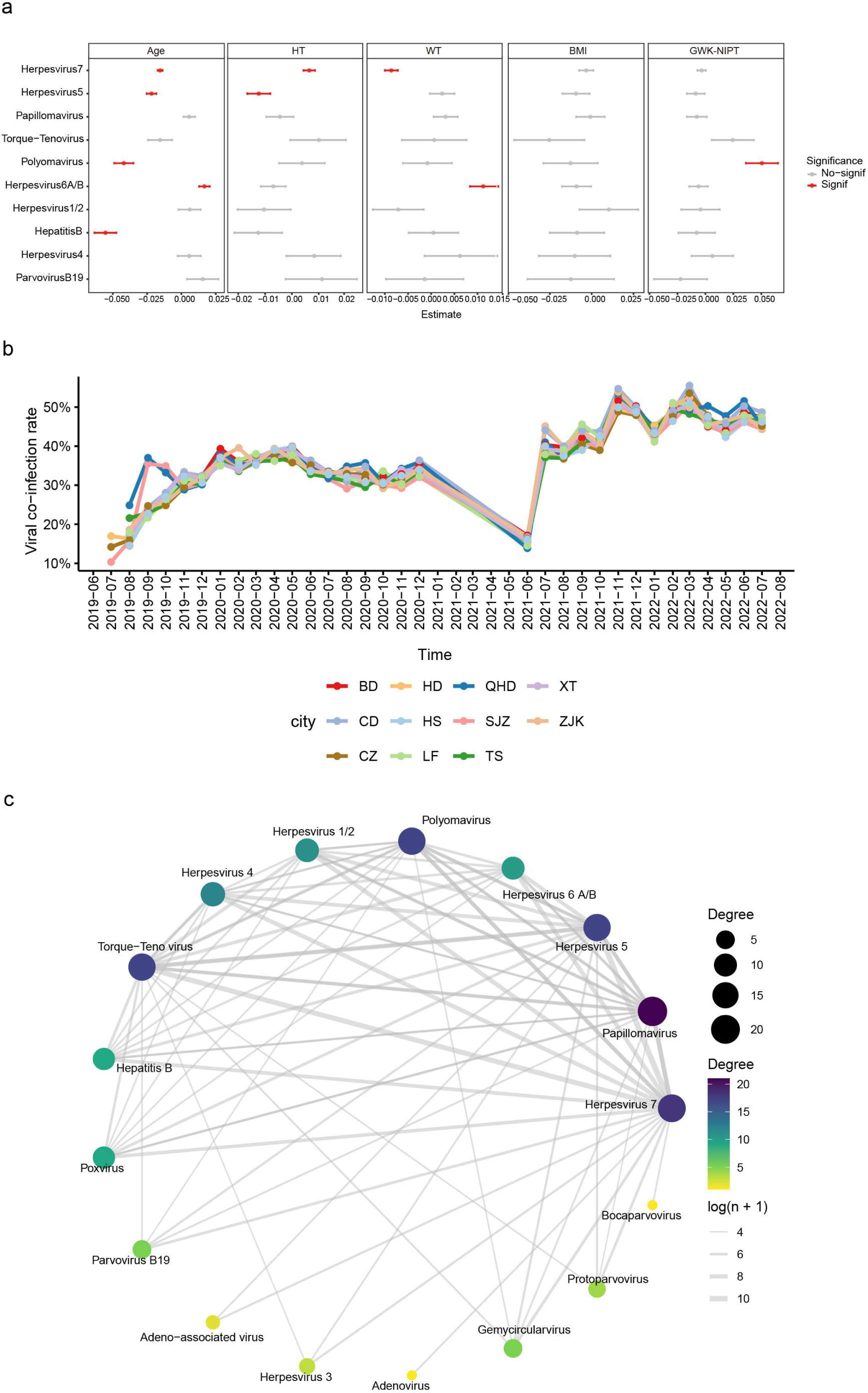
Multidimensional analysis of viral infection risk factors, regional epidemic dynamics, and viral co-infection network. **(a)** Schematic of logistic regression analysis of viral infections and maternal baseline information. The viral infection status is used as the dependent variable. Different boxes represent various pregnancy characteristics, and lines represent the 95% confidence intervals. The red line indicates a p-value < 0.05, signifying significant correlation. The raw data can be found in Supplementary Table 1**. (b)** Line chart of co-infection rate changes over time in cities in Hebei Province. **(c)** Network diagram of co-infections between different virus genera. Each node represents a virus genus, with the size of the circle indicating the frequency of co-infections for that genus. The thickness of the connecting lines represents the frequency of co-infections between two virus genera.

**S5 Fig.**
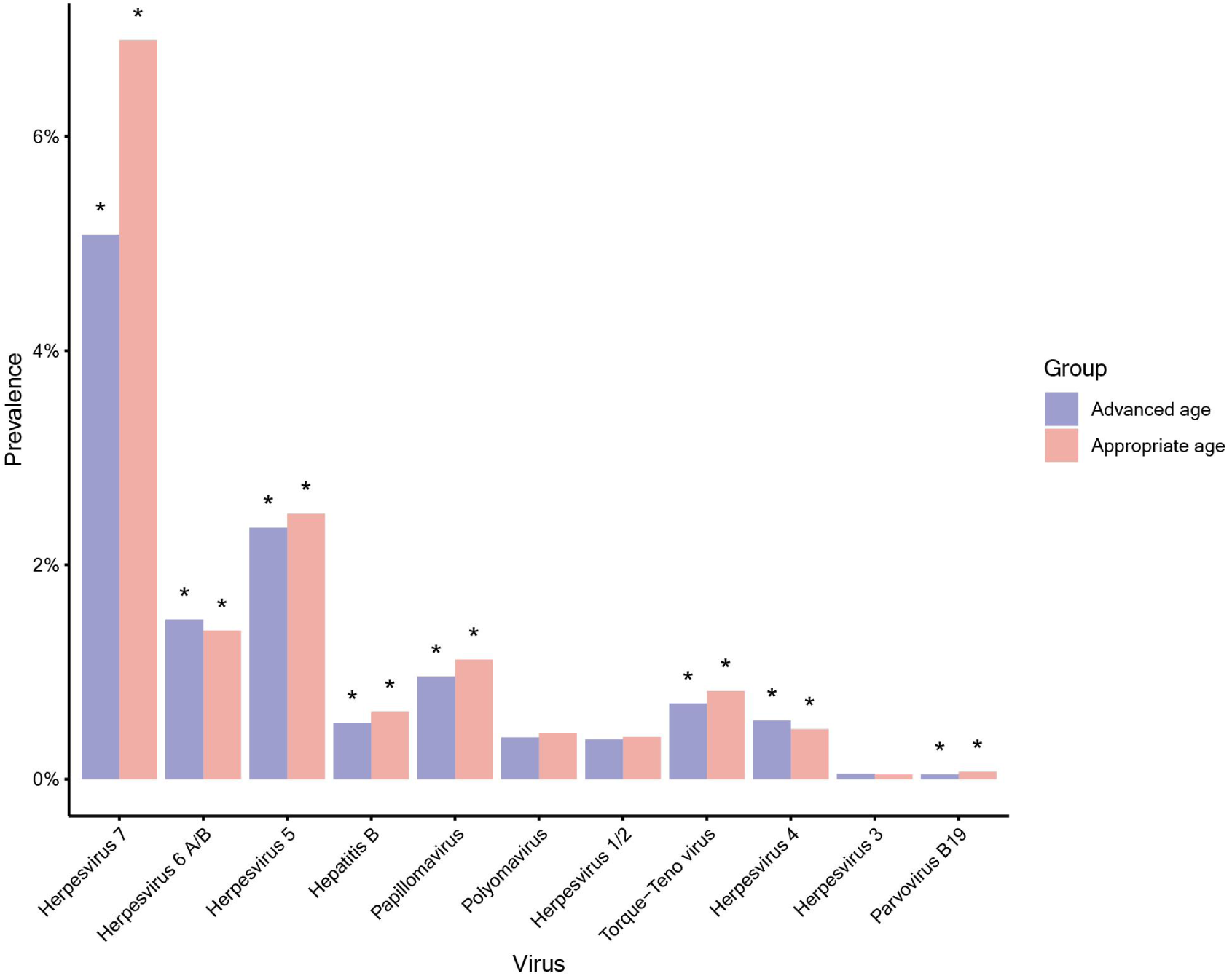
Comparison of viral prevalence between age-appropriate and advanced maternal age pregnant women. * Indicates a statistically significant difference in virus prevalence between the two groups (p < 0.05).

**S6 Fig.**
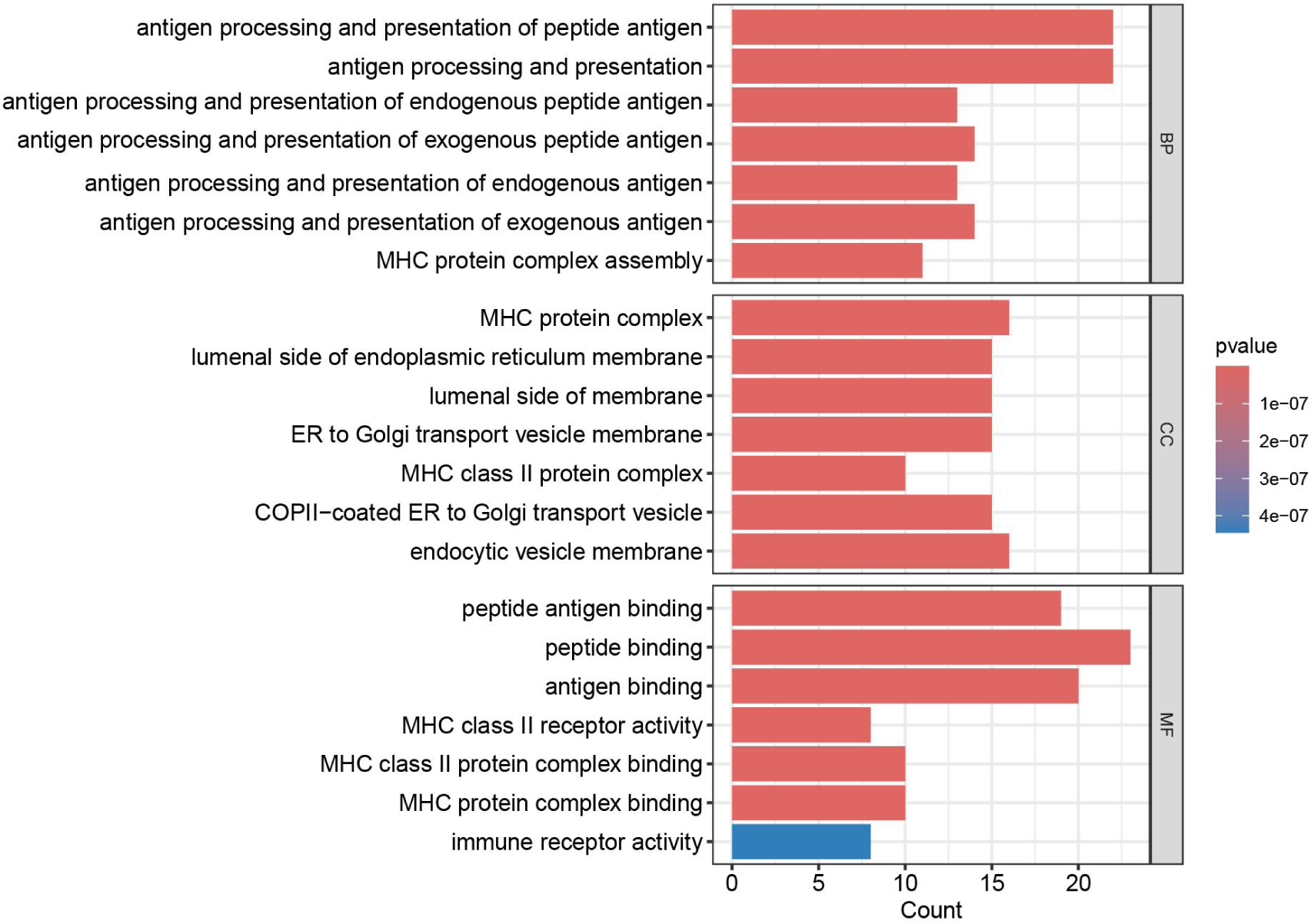
GO enrichment analysis results of susceptibility-related genes for three viruses (HHV7, HHV5, HBV). The three modules of the bar chart respectively display functional annotations at the levels of biological processes, cellular components, and molecular functions.

